# Complementary value of molecular, phenotypic and functional aging biomarkers in dementia prediction

**DOI:** 10.1101/2024.06.23.24309078

**Authors:** Andreas Engvig, Karl Trygve Kalleberg, Lars T. Westlye, Esten Høyland Leonardsen, Alzheimer’s Disease Neuroimaging Initiative

## Abstract

DNA methylation age (MA), brain age (BA), and frailty index (FI) are putative aging biomarkers linked to dementia risk. We investigated their relationship and combined potential for prediction of stages of cognitive impairment and future risk of dementia using the ADNI database. Of several MA algorithms, DunedinPACE had the strongest association with neuropsychological tests and was included alongside BA and FI in predictive analyses.

The pairwise correlations between age- and sex-adjusted measures for MA (aDundedinPACE), brain age (aBA), and frailty (aFI) were low (all <0.15). In a prediction model including age, sex, and aFI, we achieved an area under the curve (AUC) of 0.95 for differentiating cognitively normal controls (CN) from dementia patients in a held-out test set. The best models for CN vs. mild cognitive impairment (MCI) and MCI vs. dementia contained age, sex, aFI and aBA as predictors, and achieved out-of-sample AUCs of 0.82 and 0.83 respectively. When combined with clinical biomarkers (apolipoprotein E ε4 allele count, memory, executive function), a model including aBA and aFI predicted 5-year dementia risk among MCI patients with an out-of-sample AUC of 0.83. aDunedinPACE did not improve the predictions. FI had a stronger adverse effect on prognosis in males, while BA’s impact was greater in females.

Our findings highlight complementary value of BA and FI in dementia prediction. The results support a multidimensional view of dementia, including an intertwined relationship between the biomarkers, sex, and prognosis. The tested MA’s limited contribution suggests caution in their use for individual risk assessment of dementia.

## Introduction

Dementia prevalence increases exponentially after age 65 [31], but the underlying biological mechanisms linking aging to dementia remain elusive [12]. One hypothesis holds that neurodegenerative diseases leading to dementia may be manifestations of accelerated aging [16]. By quantifying deviations in biological age from expected chronological age with aging biomarkers, we might identify individuals at higher dementia risk and assess the effects of interventions targeting aging and aging-related neurodegenerative processes [12, 16].

Various biomarkers of biological age or aging have shown promise in predicting a diagnosis of dementia or future risk of the disease [3, 6, 28, 29, 35–37, 40, 42, 46]. Still, geroscience lacks a standard definition of biological aging and its ideal biomarker [8, 47]. The search for biomarkers is complex due to aging manifesting at multiple levels: molecular, phenotypic, and functional [9]. Here, we investigated the interrelated role of three leading aging biomarkers in dementia risk prediction, with each biomarker representing one of the three levels proposed by Ferruci, et al. [9].

At the molecular level, alterations in methylation patterns of DNA is a hallmark of aging [24]. The methylation states in CpG dinucleotides across the genome can be measured in peripheral blood [15]. Algorithms called epigenetic clocks leverage this data to calculate an individual’s epigenetic or methylation age (MA). Often, MA is statistically adjusted for chronological age to generate an “age acceleration” metric [14], which quantifies deviation of an individual’s methylation pattern from what is expected given their chronological age.

Here, positive values indicate that an individual’s biological age as reflected by MA is higher than their chronological age. Several studies have used epigenetic age acceleration to study dementia risk [10, 20, 26, 36, 37, 45, 48]. For instance, Mcmurran, et al., found that epigenetic age acceleration, as measured by the Horvath and GrimAge MAs increases future dementia risk [26]. Another study found that an epigenetic clock reflecting aging pace (DunedinPACE) was associated with current cognitive impairment and future dementia risk [36, 37]. Other studies, including a systematic review, have yielded mixed results [10, 45, 48] and highlight the need for further research. While single-gene methylation states were shown to predict a diagnosis of dementia due to Alzheimeŕs disease (AD) with high in-sample accuracy in one study [43], we are unaware of studies examining the predictive power of MA algorithms for individual dementia risk assessment based on out-of-sample verification. In contrast, the predictive potential of phenotypic biomarkers, in particular those captured by brain age models, has been more frequently examined in the context of individual risk assessment [11, 25, 29].

A notable example of a phenotypic aging biomarker is the difference between magnetic resonance brain imaging (MRI)-predicted and chronological age, denoted the brain age gap (BAG) [1, 11, 14]. The individual’s brain age (BA) is calculated by algorithms trained with machine learning, with recent BA models repeatedly producing reliable estimates across wide age ranges [1]. Studies have found positive associations between elevated BAG, AD biomarkers and cognitive status [28, 46]. Importantly, BAG has also been tested for individual dementia prediction. For example, using BAG, Persson, et al., achieved an area under the receiver operating characteristic curve (AUC) of 0.68 in classifying individuals with dementia from non-dementia [29].

Aging eventually manifests as functional deterioration [9, 47]. Functional aging is frequently quantified using a frailty index (FI) [33, 34]. FI is a composite score that reflects accumulation of health deficits [33]. Multiple studies have demonstrated that a greater degree of frailty, as measured by higher FI scores, correlates positively with poorer cognitive status and future dementia risk [4], even independent of cognitive test results [35]. The latter study by Song et al., [35], obtained an AUC of 0.64 and 0.66 for prediction of 5-year and 10-year future dementia risk.

A cross-sectional study by Phyo and colleagues, [30] was the first to compare the three reviewed biomarkers representing different hierarchical levels of aging (MA, BAG and FI). The authors found mostly low correlation between MA measures and FI, while BAG was not correlated with either. The findings suggested a possible complementary value of the three in predicting age-related disease risk. While this has yet to be examined in the context of dementia, studies investigating another aging-related endpoint — mortality — offer support for this notion. Li, Ploner, and colleagues [21] as well as Li, Zhang, et al. [22], found that the combination of different epigenetic clocks and FI scores enhanced mortality risk prediction compared to using any of these biomarkers in isolation. Moreover, Cole et al., found that combining BAG and MA improved mortality risk prediction [5]. These findings highlight the potential power of combining biomarkers from different levels of aging to improve prediction of age-related outcomes. To date, a direct comparison of molecular, phenotypic, and functional aging biomarkers in the context of dementia prediction is lacking.

Our primary objective was to assess the individual and combined predictive value of the reviewed aging biomarkers (measures of MA, BA, and FI) for current cognitive impairment and future dementia risk utilizing the Alzheimer’s Disease Neuroimaging Initiative (ADNI) database. We hypothesized that combining biomarkers from each of the hierarchical levels of aging would improve model performance compared to using them individually. Our secondary objective was to examine the predictive potential of the aging biomarkers when incorporated into models alongside well established, accessible clinical biomarkers: number of apolipoprotein E ε4 (APOE4) alleles and cognitive tests (memory, executive function). We anticipated that the aging biomarkers would provide added predictive value beyond these clinical markers.

For both objectives, we employed machine learning classifiers with out-of-sample verification, using AUC as our main performance metric. Theoretically, functionally apparent aging emerges when resilience mechanisms at upstream levels become exhausted. Consequently, we hypothesized that "age acceleration" at the functional level (measured by FI) would be a stronger predictor of dementia-related outcomes compared to an upstream phenotypic measure (BA). In turn, we expected phenotypic age acceleration to be a stronger predictor than upstream molecular metrics. Finally, we expected sex disparities in the aging biomarkers [30] and conducted sensitivity analyses to evaluate sex differences in their predictive power.

## Methods

### Data source

Data used in the preparation of this article were obtained from the ADNI database (adni.loni.usc.edu). ADNI was launched in 2003 as a public-private partnership, led by Principal Investigator Michael W. Weiner, MD. The primary goal of ADNI has been to test whether serial MRI, positron emission tomography, other biological markers, and clinical and neuropsychological assessment can be combined to measure the progression of mild cognitive impairment (MCI) and early AD. For up-to-date information, see www.adni-info.org. For the purposes of the present study, we drew subjects from three observational prospective case–control ADNI cohorts called ADNI1, ADNI2 and ADNIGO. Eligible subjects had available brain MRI, DNA methylation and FI data obtained within 90 days of the baseline visit; cognitive test results and demographics were drawn from the baseline visit. Data are publicly available (https://ida.loni.usc.edu/).

### Sample

Inclusion criteria included age 55 to 90 years; study partner to provide evaluation of function; speaks English; ability to undergo all testing, blood samples for genotyping and biomarkers, and neuroimaging procedures; completed six grades of education or work history; for women postmenopausal or surgically sterile, not depressed, and a modified Hachinski score less than five to rule out vascular dementia. Individuals with dementia (hereafter abbreviated “DEM”) satisfied criteria for NINCDS/ADRDA for probable AD DEM. Importantly, we used the clinical diagnosis reported in the ADNI database and did not require Alzheimer’s disease pathology biomarkers. Subjects enrolled as MCI had memory complaints verified by a study partner, Mini Mental Status Examination (MMSE) score of 24 to 30, Clinical Dementia Rating (CDR) global score (CDR-GS)[=[0.5 with sum of boxes (CDR-SB) score of at least 0.5, and general cognition and functional performance sufficiently preserved such that a diagnosis of DEM could not be made. AD biomarkers were not required for a diagnosis of MCI in the present study. Cognitively normal (CN) controls had MMSE scores of 24 to 30, CDR-GS=0 (with CDR-SB score=0) and were deemed normal based on an absence of significant impairment in cognitive functions or activities of daily living. In addition to the functional tests used to determine diagnosis, the participants underwent standard neuropsychological assessment at baseline, including Rey’s Auditory Verbal Learning Test (RAVLT) and the Trail Making Test (TMT) which probes hallmark cognitive subdomains (memory and executive function, respectively) associated with AD DEM [7, 19, 32, 44]. For the present study, we selected the immediate recall part of RAVLT (simply denoted RAVLT throughout for brevity) for memory and part B of TMT (TMT-B) for executive function.

### Molecular age

We quantified biological age at the molecular level by evaluation of DNA methylation (DNAm) patterns in white blood cells obtained from peripheral blood samples. DNAm data profiling was previously performed by the ADNI investigators for 653 unique ADNI participants using the Illumina Infinium HumanMethylationEPIC BeadChip Array (www.illumina.com). We used a subset of these (n=385) that also had available MRI and FI data within 90 days of the baseline visit. If more than one DNAm sample existed for each unique participant, we selected the sample from the baseline visit. If a baseline sample was not available, we drew the sample at the temporally closest time point to baseline, excluding cases where were the baseline to sample interval was 90 days or more apart. Median (absolute) time between baseline visit and blood sampling was 0.5 days (interquartile range (IQR) = 0 to 7) and was similar between diagnostic groups and stable versus progression MCI (Wilcoxon tests, p-values > .05).

Several state-of-the-art MA algorithms may be used to obtain measures of aging from blood cell DNAm data. Here, we selected five candidate MA algorithms that have been associated with either neuropsychological test results, diagnosis and/or future conversion to dementia risk in one or more observational studies [15, 26, 27, 37, 45, 48]: Horvath’s first-generation epigenetic clock (DNAmAge), Horvath’s epigenetic “Skin-Blood” clock (Skin-BloodClock), second generation PhenoAge and GrimAge epigenetic clocks, and a third generation "pace of aging" MA measure, DunedinPACE. Given the lack of a single or gold standard MA algorithm for DEM prediction and the inconsistent association between present MA measures and neurocognitive outcomes, the best subset of MA metrics was decided via exploratory data analysis in the training set prior to the predictive modelling (details provided below).

### Phenotypic age

We used BA to operationalize aging at the phenotypic level. We estimated BA using a previously published deep neural network that has been shown to generalize to unseen scanners and cohorts [18], and which is freely available online (https://github.com/estenhl/pyment-public). Briefly, this is a convolutional neural network with an architecture consisting of six blocks of convolutional and max-pooling layers. Prior to modelling, T1-weighted MRI scans were minimally processed to remove non-brain tissue with the recon-all pipeline from FreeSurfer 5.3. Further, they were rigidly registered to the same stereotactic space, using flirt from FSL with six degrees of freedom. Further details of the pipeline are described in the original publication [18]. In cases where participants had multiple MRI scans, we chose the imaging session closest to baseline and excluded cases where were the baseline to scan interval was 90 days or more apart. The median (absolute) baseline to scan interval was 16 days (IQR = 10 to 27).

### Functional age

We operationalized aging at a functional level using a 26-item FI based on the accumulation of deficits model of frailty by Rockwood and Mitnitski [33]. We manually selected health deficits available at the screening or baseline visit in the ADNI database following a standard procedure [34]. Then, we used a combination of factor analysis of mixed data, cluster analysis, and regression analysis to reduce the number of deficits and maximize explained variance (for details regarding calculation, included items and cut-offs, see [6]). The code for calculating the present FI using ADNI data is also freely available (https://github.com/LAMaglan/ADNI-FI-clustering). The FI variable contained health deficits covering a range of systems including non-cognitive clinical tests, such as blood test results (red and white blood cells counts), blood pressure, history of disease, symptoms such as low energy, alterations in gait and functional measures. Higher FI scores indicate greater degree of frailty (i.e., lower level of physical or systems-level function). The present FI has been validated against mortality and DEM risk in ADNI previously [6].

### Age and sex-adjusted biomarkers

To derive comparable measures of aging at the three different levels, we first adjusted each measure by regressing out sex and chronological age using linear models. These were fit using the training data (see below), and then applied to both the training and test data to produce residuals instead of raw measures, representing sex and age-adjusted values. Next, we standardized the residuals by subtracting the mean and dividing by the standard deviation to obtain z-scores for subsequent modelling. The deviations from expected values based on sex and chronological age at different levels (i.e., biological, phenotypic, and functional) was denoted by adding the prefix ’a’ to the adjusted measures: aDunedinPACE, aBA, and aFI. The same age-adjustment and standardization procedure was done for RAVLT (aRAVLT) and TMT-B (aTMT-B) to render variants of these uncorrelated with sex and age. To explicitly assess the impact of chronological age and sex in the predictive models, we included these as predictors when appropriate.

### Statistical analyses

Before any analyses were performed, we split the data into subsets to facilitate hyperparameter-tuning and unbiased out-of-sample estimates of model performance (Fig. 1). We aimed to use as much data as possible to fit the models, despite the participants having various combinations of the relevant measures. Starting from the full dataset (n=1876), we first extracted all participants lacking methylation data into a stage 1 modelling dataset (n=1491), to be used for identifying the optimal selection of BA and FI (e.g., only aBA, only aFI, or aBA+aFI) as predictors, across all predictive tasks. The remaining 385 participants (with DNAm data) were stratified using diagnosis, age, and sex, before 260 were drawn to form the dataset for a second modelling stage (stage 2 dataset). The remaining 125 participants were reserved in a held-out test set.

**Fig. 1.**
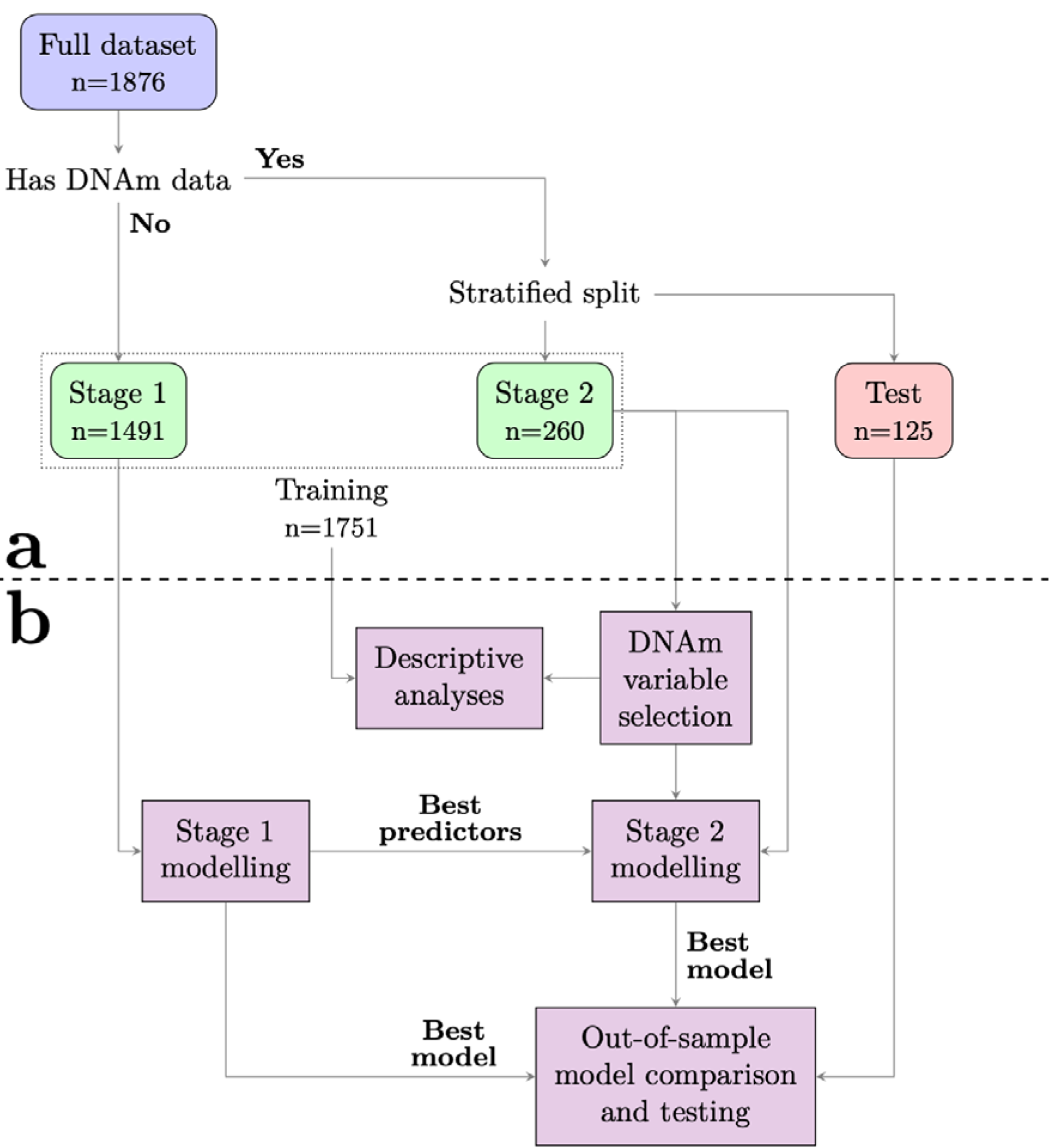
**a)** Data were split into subsets to facilitate hyperparameter-tuning and obtain unbiased out-of-sample estimates of model performance. Due to various combinations of missing biomarker data, we first extracted all participants with complete frailty index (FI) and brain age (BA) data, but lacking methylation data into a stage 1 modelling training set (n=1491). This was used to identify the optimal selection of age- and sex-adjusted BA and FI (i.e., aBA, aFI, or aBA+aFI) as predictors across all predictive tasks. The remaining 385 participants (with complete FI, BA and DNA methylation data) were stratified using diagnosis, age, and sex and divided into another training set for a second modelling stage (stage 2) or a hold-out test set. **b)** All training data was utilized for descriptive statistics. We employed data-driven variable selection to determine which DNA methylation age markers to use. After selecting the best combination of BA and FI as predictors, stage 2 modeling was done to find the optimal selection of all three types of aging biomarkers. The best models from stage 1 and 2 were compared in a cross-validation loop, before evaluating the best model in the test set to obtain an unbiased estimate of model performance

Given the lack of a principal MA measure of molecular age, we performed data-driven feature selection in the stage 2 training dataset to identify a subset of candidate MA measures prior to any modelling. Here, we tested for associations between each of the five adjusted MA measures (aDNAmAge, aGrimAge, aPhenoAge, aSkin-BloodClock, aDunedinPACE) and age- and sex-adjusted tests of memory (aRAVLT) and executive function (aTMT-B), which are well-known risk factors for DEM [7, 44]. We employed linear regression models for each univariate analysis. Next, we retrieved the coefficient and p-value for each of these bivariate relationships from the models and corrected the p-values for multiple tests to reduce the false discovery rate (FDR) using the Benjamini-Hochberg procedure. Finally, MA measures with at least one p-value <0.05 post-correction for any of the predictive targets were retained for the subsequent analyses.

Next, we performed multiple descriptive analyses in the training set to assess the interrelationship between the aging biomarkers, and how they relate to the diagnostic and prognostic groups. Here, for prognostic comparison, we divided MCI subjects into those who converted to DEM within 5-years from baseline (progressive MCI, pMCI) and those who remained stable (stable MCI, sMCI). First, we investigated the distribution of values for each measure in the two groups comprising each of four binary partitions of the training set (Fig. 3a): CN vs DEM patients, CN vs MCI patients, DEM vs MCI patients, and sMCI vs pMCI, respectively. For each comparison, a Kruskal-Wallis H-test was employed to determine whether the medians of the two groups differed significantly, with p-values corrected to control the FDR. Lastly, we investigated the covariance between the measures, also in the stage 2 dataset, by computing all pairwise Pearson correlations (Fig. 3b). Here we also included their correlation with aTMT-B, aRAVLT and the number of APOE4 alleles (0, 1 or 2), coded as an ordinal variable.

We then fit logistic regression models with an l_l_-loss to assess the predictive value of the aging biomarkers. We first tested diagnostic predictions: For each of three predictive tasks (CN vs DEM, CN vs MCI, MCI vs DEM) this was performed in two stages, based on the stage 1 and 2 datasets. First, we used stage 1 data to perform a ten-fold cross-validation (CV) to determine the best combination of aBA and aFI and the optimal value for the regularization parameter A, fitting multiple models for each permutation of these two. Each of these models included age and sex as additional predictors, and we also fit a fourth model containing only age and sex as a baseline. The best stage 1 model was determined as the one yielding the highest mean AUC across the ten validations. Next, we performed a similar ten-fold cross-validation (CV) using the stage 2 data to determine whether to include aDunedinPACE, and alternatively the ideal value of A. Here, we fit two models for aDunedinPACE: one containing only age and sex as additional predictors, and one also including the best stage 1 predictors. Also, here we selected the model yielding the highest mean CV AUC. Finally, we calculated the AUC of the best performing model in the held-out test set. We performed this process to determine the optimal combination of predictors among the three aging biomarkers, and to procure an unbiased out-of-sample estimate of predictive performance.

In the final analysis we investigated whether any of the three aging biomarkers at baseline provided prognostic value for detecting which MCI patients would progress into DEM within 5 years, by differentiating the sMCI and pMCI cohorts. We deemed a 5-year timeframe to be clinically relevant while ensuring enough converting subjects for modelling. First, we performed the same variable selection process as above to identify the best set of predictors among the aging biomarkers. Next, we fit a model using clinical biomarkers, e.g. aTMT-B, aRAVLT and APOE4, using the same training data as the candidate model chosen for the aging biomarkers (e.g. either stage 1 or stage 2 data). Finally, we fit a model containing both the best set of aging biomarkers measures and the clinical (also using the same data). To investigate whether the two sets of covariates (aging, clinical) were complementary, we triangulated the AUCs achieved in the test set between these three models. For interpretability, and to allow for comparisons with other studies, we calculated the balanced accuracy, sensitivity, specificity and positive predictive value of the best performing model alongside the AUC. Given the l_l_-loss employed attempting to nullify the coefficients of predictors not contributing to the predictions, we also report the non-zero coefficients used in the best performing model as a proxy of which variables contributed to the prediction.

To tease apart potential sex differences in the interrelatedness and predictive power of the proposed biomarkers, we supplemented the main prediction analyses with a similar analysis while splitting the dataset based on sex. Here, we fit models independently for males and females, keeping the standardized aging biomarkers and chronological age as predictors. Due to the smaller sample size resulting from analyzing male and female sub-groups separately, we compared the sex-specific results using 100 bootstrapped training and test subsets, to yield distributions of both performance metrics and coefficient estimates.

## Results

### Descriptive statistics

The training dataset consisted of 1751 subjects (Fig. 1). As described under Methods, a subset of 260 subjects had DNAm data obtained within 3 months of baseline and was used for stage 2 modeling. Within this subset, 158 had a baseline diagnosis of MCI with complete aging biomarker data where 22% converted to DEM within 5 years (Table 1).

**Table 1.**
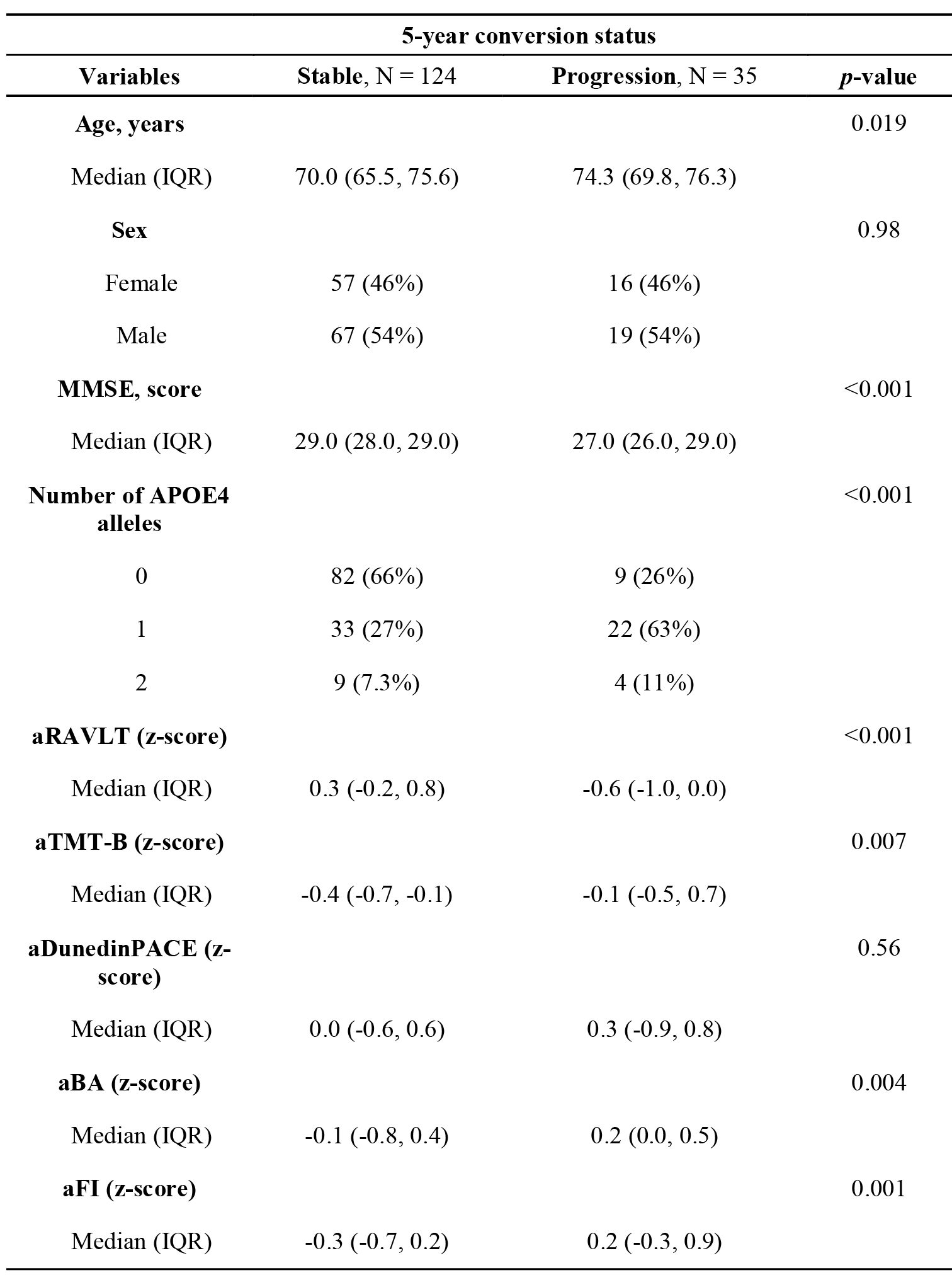
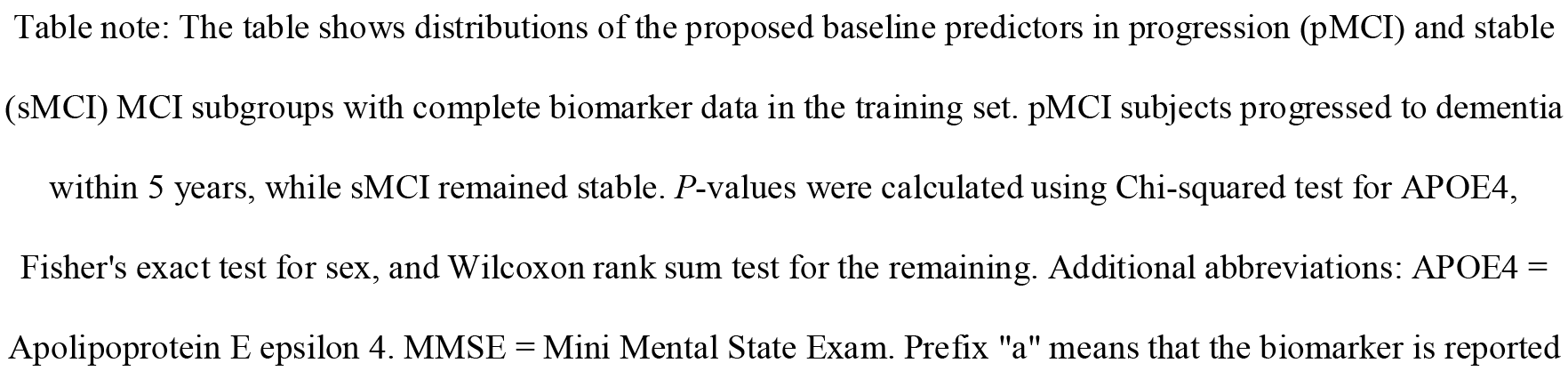
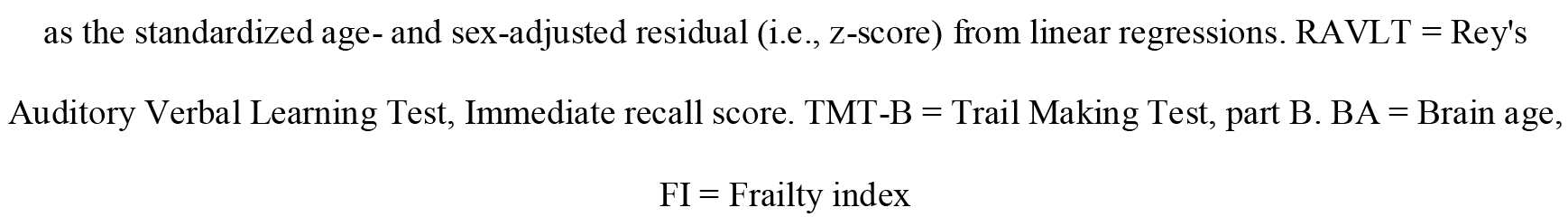
Baseline characteristics of mild cognitive impairment (MCI) subgroups in training sample with complete biomarker data (N = 159)

| 5-year conversion status |  |  |  |
| --- | --- | --- | --- |
| Variables | Stable, N = 124 | Progression, N = 35 | *p*-value |
| **Age, years** |  |  | 0.019 |
| Median (IQR) | 70.0 (65.5, 75.6) | 74.3 (69.8, 76.3) |  |
| **Sex** |  |  | 0.98 |
| Female | 57 (46%) | 16 (46%) |  |
| Male | 67 (54%) | 19 (54%) |  |
| **MMSE, score** |  |  | <0.001 |
| Median (IQR) | 29.0 (28.0, 29.0) | 27.0 (26.0, 29.0) |  |
| **Number of APOE4 alleles** |  |  | <0.001 |
| 0 | 82 (66%) | 9 (26%) |  |
| 1 | 33 (27%) | 22 (63%) |  |
| 2 | 9 (7.3%) | 4 (11%) |  |
| **aRAVLT (z-score)** |  |  | <0.001 |
| Median (IQR) | 0.3 (-0.2, 0.8) | -0.6 (-1.0, 0.0) |  |
| **aTMT-B (z-score)** |  |  | 0.007 |
| Median (IQR) | -0.4 (-0.7, -0.1) | -0.1 (-0.5, 0.7) |  |
| **aDunedinPACE (z-score)** |  |  | 0.56 |
| Median (IQR) | 0.0 (-0.6, 0.6) | 0.3 (-0.9, 0.8) |  |
| **aBA (z-score)** |  |  | 0.004 |
| Median (IQR) | -0.1 (-0.8, 0.4) | 0.2 (0.0, 0.5) |  |
| **aFI (z-score)** |  |  | 0.001 |
| Median (IQR) | -0.3 (-0.7, 0.2) | 0.2 (-0.3, 0.9) |  |

Table note: The table shows distributions of the proposed baseline predictors in progression (pMCI) and stable (sMCI) MCI subgroups with complete biomarker data in the training set. pMCI subjects progressed to dementia within 5 years, while sMCI remained stable. *P*-values were calculated using Chi-squared test for APOE4, Fisher's exact test for sex, and Wilcoxon rank sum test for the remaining. Additional abbreviations: APOE4 = Apolipoprotein E epsilon 4. MMSE = Mini Mental State Exam. Prefix "a" means that the biomarker is reported
as the standardized age- and sex-adjusted residual (i.e., z-score) from linear regressions. RAVLT = Rey's Auditory Verbal Learning Test, Immediate recall score. TMT-B = Trail Making Test, part B. BA = Brain age, FI = Frailty index

Correlations between adjusted MA measures and tests of memory and executive function are shown in Fig. 2. After p-value correction only two of the 15 correlations remained significant (adjusted *p*-value < 0.05): Increased aDunedinPACE was significantly associated with poorer memory, i.e., lower scores on aRAVLT ( =-0.2, adjusted *p*-value = 0.02), and worse executive function, i.e., increased scores on TMT-B ( =0.18, adjusted *p*-value = 0.03). Based on results suggesting poorer cognitive test scores with higher aDunedinPACE, we retained this as the sole MA measure for subsequent analyses.

**Fig. 2.**
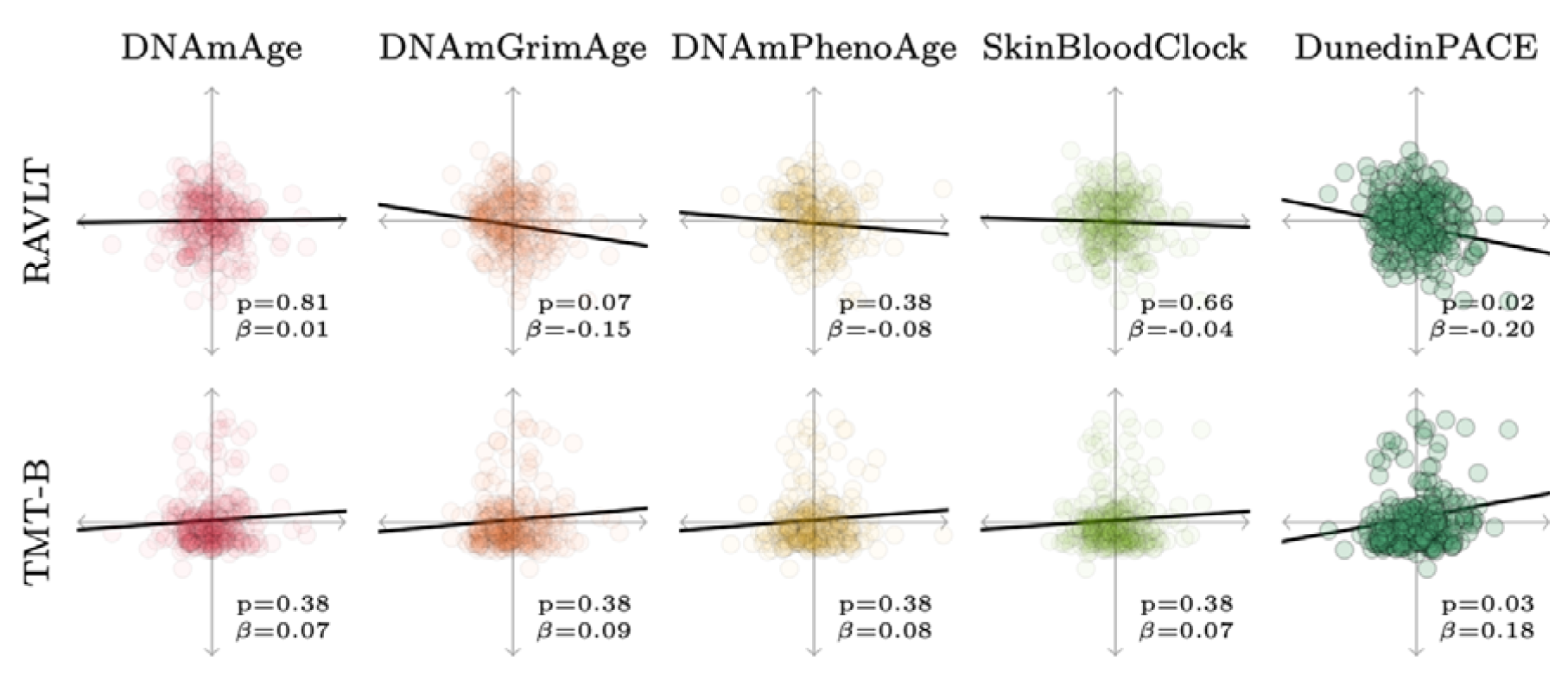
Scatterplots showing individual data and model fit lines from linear regressions from the stage 2 training dataset of age- and sex-adjusted candidate methylation ages (MA) and memory (Rey Auditory Verbal Learning Test, immediate recall (RAVLT), upper row) and executive function (Trail Making Test, part B (TMT-B), lower row). β represents the linear regression coefficient of each MA and the corresponding False Discovery Rate-corrected *p*-value

**Fig. 3.**
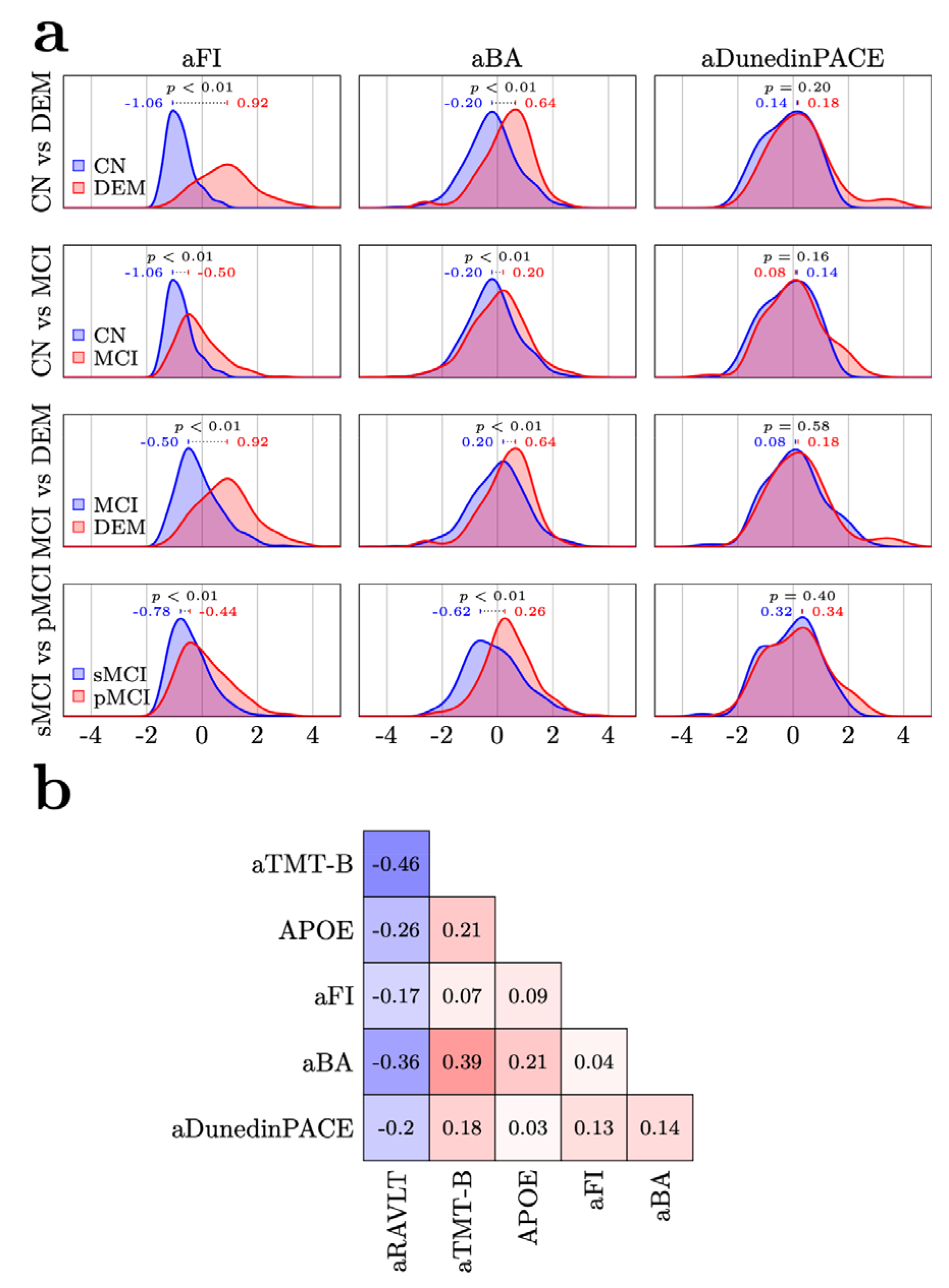
**a)** Training set density plots illustrate the distributions of age and sex-adjusted aging biomarkers (standardized residuals) for the subgroups used for diagnostic and prognostic classification. 1st row shows the distributions for subjects with normal cognition (CN) and dementia (DEM), respectively; 2nd row CN and subjects with mild cognitive impairment (MCI); 3rd row MCI and DEM. 4th row subjects with MCI progressing to DEM within 5-years (pMCI) and subjects with MCI remaining stable (sMCI). The blue and red values are subgroup adjusted aging biomarker medians. *P*-values are False Discovery Rate-adjusted from Kruskal-Wallis H-tests of subgroup differences in the biomarkers. **b)** Pearson correlations between the adjusted aging biomarkers and common clinical markers in the training set. Additional abbreviations: prefix a = standardized age- and sex-adjusted residual. APOE = Apolipoprotein E epsilon 4 allele count. BA = Brain age. FI = Frailty index. RAVLT = Rey Auditory Verbal Learning Test, immediate recall. TMT-B = Trail Making Test, part B

Next, we compared distributions of the adjusted aging biomarkers in four binary partitions, each consisting of two disjunct diagnostic groups (CN vs MCI, CN vs DEM, MCI vs DEM, sMCI vs pMCI), in the training set (Fig. 3a). Here, aDunedinPACE did not show significant group differences in either comparison (all adjusted *p*-values > 0.05), while both aBA and aFI showed significant differences between the groups in all four (all adjusted *p*-values < 0.01).

For aBA, the largest group difference was observed between the sMCI and pMCI groups (0.88 difference in group medians), whereas for aFI the largest discrepancy was seen for the CN and DEM patients (1.98 difference in group medians).

Finally, we computed bivariate correlations for the adjusted aging biomarkers, executive and memory function, and APOE4-status, still in the training sample (Fig. 3b). Besides aRAVLT which is coded in the opposite direction (i.e., lower values mean more severe impairment), all measures were positively correlated, indicating general agreement. The highest absolute correlation was observed between aTMT-B and aRAVLT (-0.46). The highest correlation containing at least one aging biomarker was between aBA and aTMT-B (0.39). Among the aging biomarkers, the highest correlation was observed between aDunedinPACE and aBA (0.14). Although there was concordance in the directionality among the three aging biomarkers, their correlation was relatively low (all <0.15), underscoring the possibility that they are sensitive to complementary information. Correlations among age-adjusted biomarkers for males and females separately are shown in Supplemental Fig. 1.

### Predictive analyses: BA and FI provide complementary information for predicting current cognitive status

Next, we performed predictive analyses, utilizing the adjusted aging biomarkers as predictors in three binary diagnostic classification problems: CN vs DEM, CN vs MCI and DEM vs MCI (Fig. 4).

**Fig. 4.**
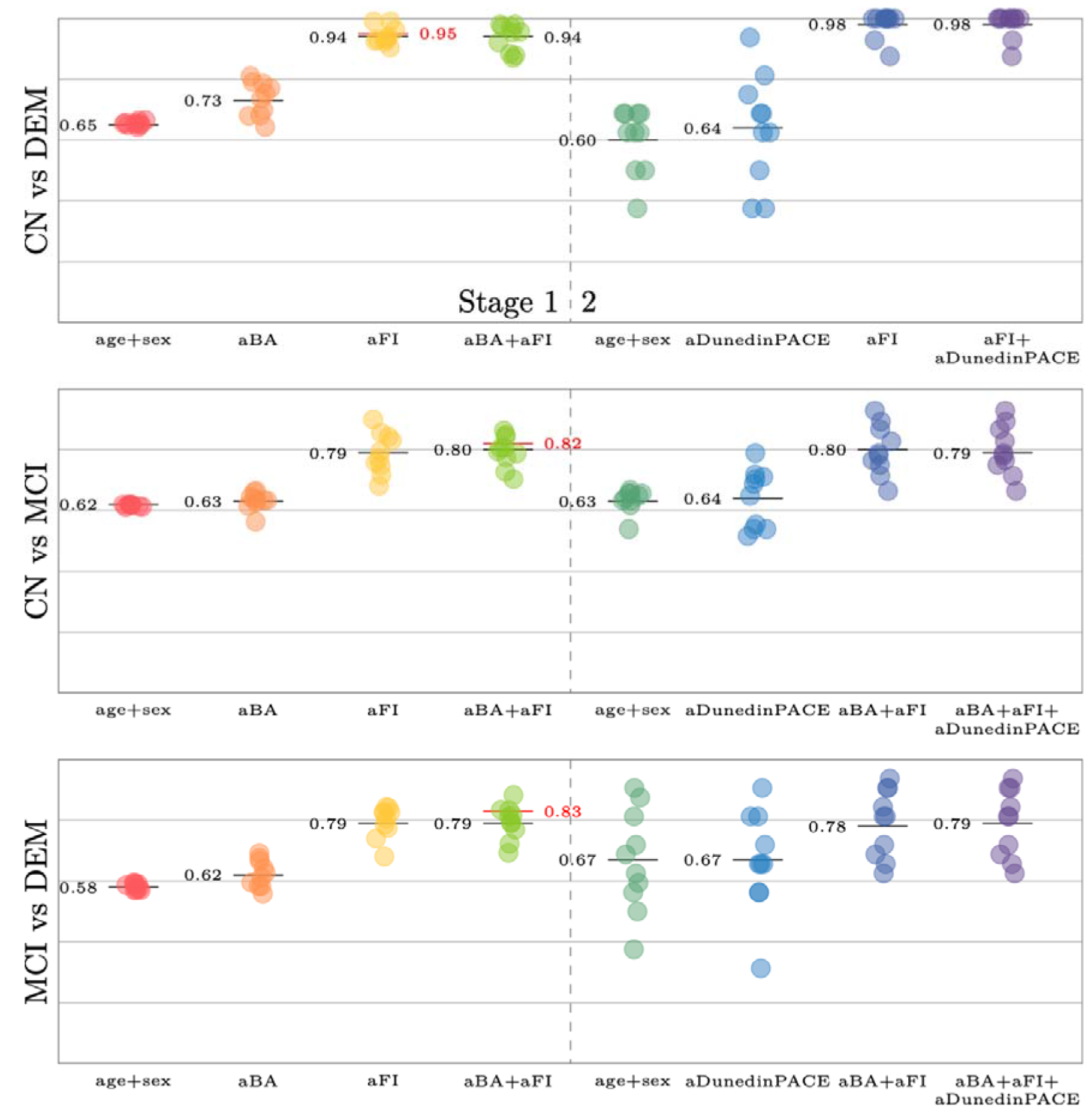
Predictive performance––reported as Area under the Receiver Operating Characteristic Curve (AUC)–– for the various models for each of three predictive (diagnostic) tasks. For each task, a baseline model was first fit in the stage 1 dataset using age and sex as predictors. Next, models including standardized age- and sex-adjusted brain age (aBA) and frailty index (FI) residuals, both independently and in combination, were trained using the same data. The best model was retained for the stage 2 data and compared with another baseline-model, a model containing aDunedinPACE, and a model adding aDunedinPACE to the best models from stage 1. The x-axis denotes these different models, and the y-axis denotes AUC. Individual points represent performance in independent folds in the cross-validation, whereas the black line denotes their mean. The red line represents the performance of the best model in the hold-out test set. Additional abbreviations: Additional abbreviations: CN = Cognitively normal, MCI = Mild Cognitive Impairment, DEM = Dementia

For CN vs DEM, the model containing only aBA (alongside age and sex) outperformed a baseline model containing only age and sex (mean CV AUC = 0.73 vs 0.65). However, this was greatly surpassed by the model containing aFI (CV AUC = 0.94), which also outperformed the one combining aBA and aFI (CV AUC = 0.94). In the stage 2 dataset the model with aFI (i.e., the best model from stage 1) reached a mean CV AUC of 0.98. This result was unchanged when adding aDunedinPACE as a predictor, indicating that neither this variable, nor aBA complemented aFI for distinguishing CN and DEM. As a final test of model efficacy, the best model (containing only aFI) achieved an out-of-sample AUC of 0.95 in the held-out test set.

For classification of CN vs MCI, the best stage 1 model contained both aFI and aBA (CV AUC = 0.80), which marginally outperformed the model with only aFI (CV AUC = 0.79). As above, the best model from stage 1 (aBI+aFA, stage 2 CV AUC = 0.80) was superior to the best stage 2 model including aDunedinPACE (CV AUC = 0.79), demonstrating its redundancy also here. For this task, the best model containing both aFI and aBA achieved an out-of-sample AUC of 0.82 in the held-out test set, indicating good classification performance.

For MCI vs DEM the best stage 1 model also included both aFI and aBA, although here the gap to the model containing only aFI was minute (AUC_aFI+aBA_ = 0.7947 vs AUC_aFI_ = 0.7946). The model containing aFI and aBA as predictors also outperformed the models including aDunedinPACE (mean CV AUC = 0.79 vs 0.78 in the stage 2 data) and achieved an AUC of 0.83 in the held-out test set.

To summarize, the best model for differentiating CN and DEM included aFI (alongside age and sex) and reached an AUC of 0.95 in the held-out test set. This indicates excellent ability to distinguish subjects with normal cognition from those living with DEM, also when applied to unseen data. For both CN vs MCI and MCI vs DEM, the best model contained aFI and aBA, yielding out-of-sample AUCs of 0.82 and 0.83 respectively. The latter results indicate that a model including age, sex, aBA and aFI had good ability to distinguish MCI from those with DEM and from those with normal cognition (CN). In supplementary analyses excluding aFI, we found that combining aBA and aDunedinPACE did not improve any of the predictions beyond using each predictor independently (Supplementary Fig. 2).

### BA and FI complement common clinical markers to predict prognosis

Next, we investigated the efficacy of the aging biomarkers to differentiate sMCI and pMCI patients, effectively attempting to answer the clinical question “Will this patient with MCI convert to DEM over the next 5 years?”. Here, the best stage 1 model included both aFI and aBA (CV AUC = 0.74, Fig. 5a), with both variables bringing substantial and similar predictive gains (CV AUC = 0.66 for only aBA and 0.71 for only aFI).

**Fig. 5.**
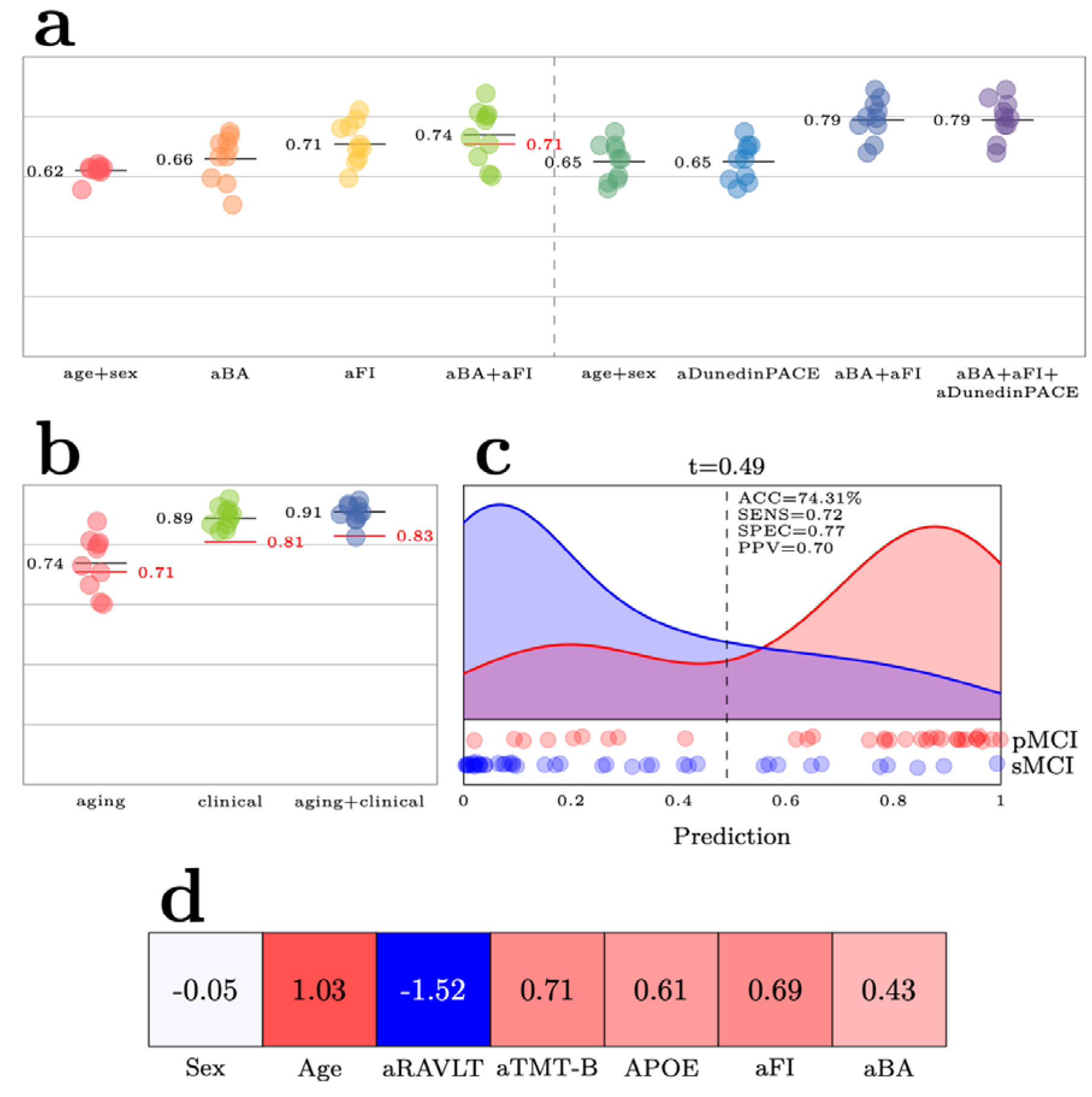
**a)** Comparison of the different prognostic models for predicting progression from mild cognitive impairment (MCI) at baseline to dementia (DEM) within 5-years. **b)** Comparison of the best prognostic aging model with a prognostic model containing tests commonly used in clinical workup of suspected DEM: Apolipoprotein E epsilon 4 allele count (APOE), the Rey Auditory Verbal Learning Test, immediate recall (RAVLT), and the Trail Making Test, part B (TMT-B). The final model denoted aging+clinical combines predictors from these two. All models included age and sex. The individual points denote Area under the Receiver Operating Characteristic Curve (AUC) in independent folds in the cross-validation (CV), the black line the CV mean, and the red line model performances in the hold-out test set. **c)** Distribution of predictions in the two groups (stable MCI, progressive MCI). The dashed line denotes the classification threshold t, determined via the receiver operating curve. **d)** The regression coefficients of the best performing model for predicting 5- year DEM progression

The models containing aDunedinPACE did not yield further predictive gains in the stage 2 CV (AUC = 0.78 vs 0.79 when only using aBA and aFI), and the model with aFI and aBA was retained, reaching an AUC of 0.71 in the held-out test set (Fig. 5b).

To establish a baseline for determining whether the aging biomarkers provide additional prognostic value beyond predictors commonly used in DEM work-up, we fitted a model using age, sex and clinical test results (aRAVLT, aTMT-B and APOE4 allele count), yielding a mean CV AUC of 0.89. Building on this, we fitted another model that included the clinical tests, age, sex, and aFI and aBA as aging biomarkers, yielding a mean CV AUC of 0.91, indicating excellent classification performance. The result suggests that the enhanced model incorporating the aging biomarkers outperformed the one with clinical markers, age and sex. This final model reached an AUC of 0.83 in the held-out test set (compared to 0.81 for the model containing only the clinical variables), suggesting good to excellent out-of-sample capability to differentiate between subjects progressing to DEM and those with a stable condition. In the test set, the enhanced model had a balanced accuracy of 74%, sensitivity of 0.72, specificity of 0.77, and positive predictive value of 0.70 (Fig. 5c). Despite the l_l_-loss attempting to eliminate superfluous covariates, all coefficients remained non-zero (Fig. 5d), with aRAVLT having the largest absolute value (-1.52), followed by age (1.03), aTMT-B (0.71), aFI (0.69), APOE4 (0.61), aBA (0.43), and sex (-0.05).

### The predictive power of aging biomarkers differs between males and females

Finally, we compared the best performing models for females and males separately. Table 2 displays the standardized regression coefficients, model performance and *p*-values for statistical tests of sex differences in coefficients and performance.

**Table 2.**
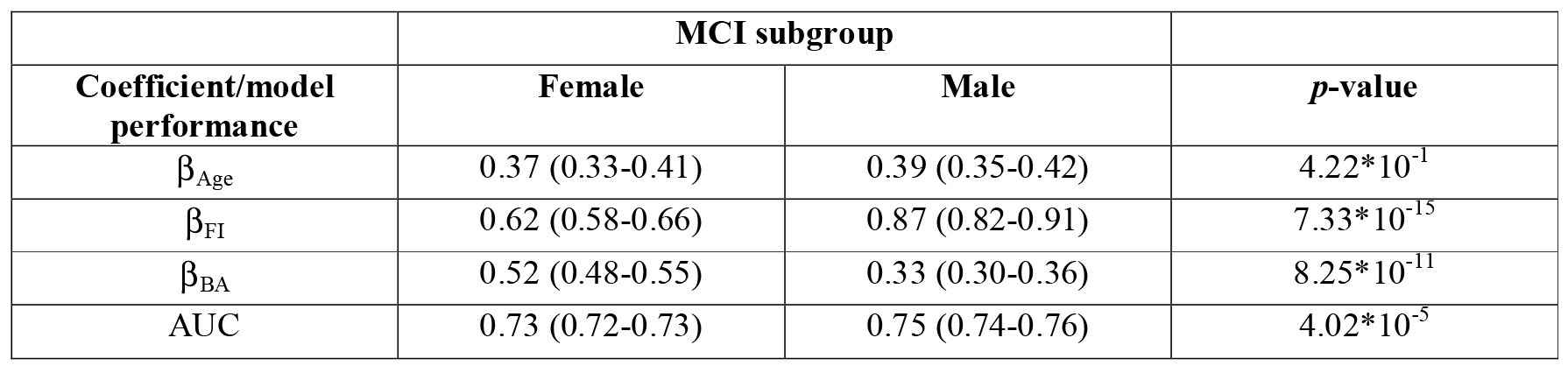
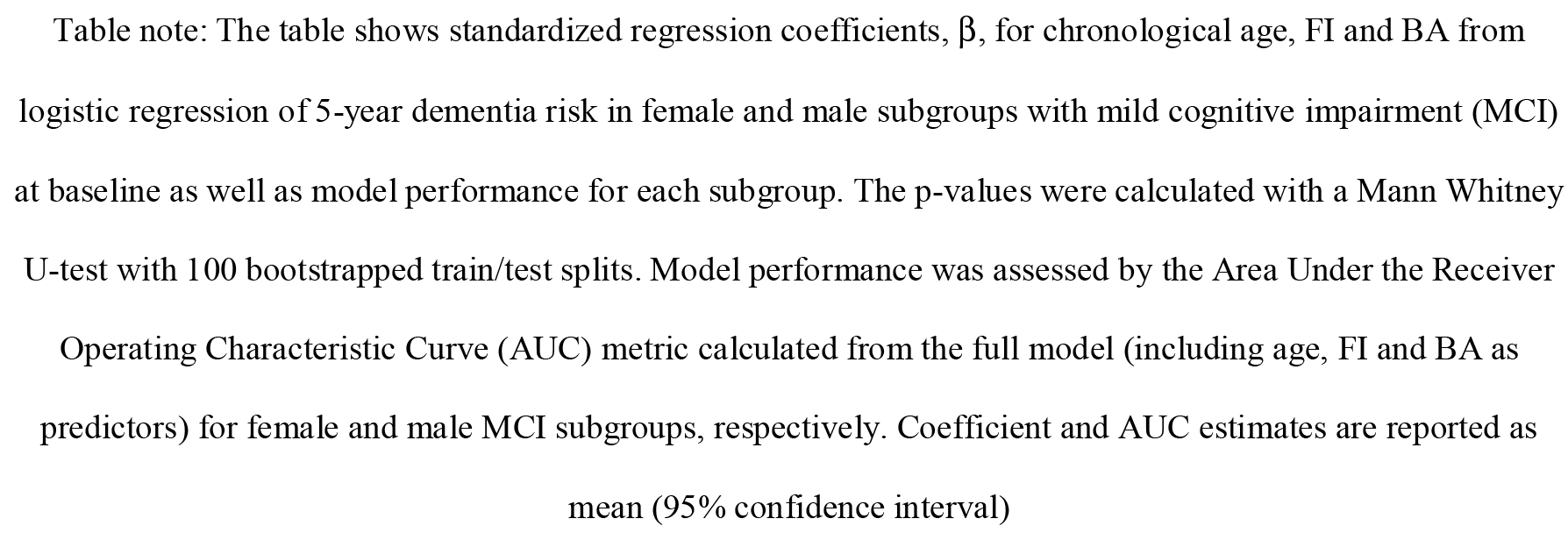
Regression coefficients and model performance for prediction of 5-year dementia conversion in female and male subgroups with baseline mild cognitive impairment.

| **Coefficient/model performance** | **MCI subgroup** |  | ***p*-value** |
| --- | --- | --- | --- |
|  | **Female** | **Male** |  |
| $\beta_{\text{Age}}$ | 0.37 (0.33-0.41) | 0.39 (0.35-0.42) | $4.22 \times 10^{-1}$ |
| $\beta_{\text{FI}}$ | 0.62 (0.58-0.66) | 0.87 (0.82-0.91) | $7.33 \times 10^{-15}$ |
| $\beta_{\text{BA}}$ | 0.52 (0.48-0.55) | 0.33 (0.30-0.36) | $8.25 \times 10^{-11}$ |
| AUC | 0.73 (0.72-0.73) | 0.75 (0.74-0.76) | $4.02 \times 10^{-5}$ |

Table note: The table shows standardized regression coefficients, $\beta$ , for chronological age, FI and BA from logistic regression of 5-year dementia risk in female and male subgroups with mild cognitive impairment (MCI) at baseline as well as model performance for each subgroup. The *p*-values were calculated with a Mann Whitney U-test with 100 bootstrapped train/test splits. Model performance was assessed by the Area Under the Receiver Operating Characteristic Curve (AUC) metric calculated from the full model (including age, FI and BA as predictors) for female and male MCI subgroups, respectively. Coefficient and AUC estimates are reported as mean (95% confidence interval)

Here, the best model of aging biomarkers predicting 5-year DEM risk (sMCI vs. pMCI) performed slightly better for males compared with females (Table 2). The standardized β- value for aFI was significantly higher in males (95% CI β 0.82 to 0.91) compared with females (β 0.58 to 0.66), clearly suggesting a greater risk of DEM progression in men with increasing level of frailty. Conversely, in the same model, increased aBA (i.e., higher apparent brain age)) was a stronger predictor of future DEM risk in females compared with males (Table 2).

The sex difference in aBA on future DEM risk was diminished in the model also including clinical biomarkers. The model including both aging and clinical biomarkers showed comparable overall performance in predicting 5-year DEM risk for males and females (AUC 0.87 and 0.86 for females and males, respectively). The sex differences in aFI β-values, however, remained significant (Table 2) and suggested higher adverse effect of increased frailty in males compared with females also when adjusting for APOE4 allele count and neuropsychological test performance.

## Discussion

We investigated the predictive value of three aging biomarkers for assessing baseline cognitive impairment and future DEM risk in ADNI. Our main findings include: 1) Age, sex and FI effectively distinguished CN from baseline DEM (out-of-sample AUC 0.95), 2) BA and FI independently enhanced prediction of MCI diagnosis (e.g., out-of-sample AUC_DEM vs MCI_ 0.83) and 5-year DEM risk in MCI subjects (out-of-sample AUC 0.71), 3) BA and FI further added prognostic value beyond routine clinical tests, with increases in frailty having a particularly strong adverse influence on males, and 4) DunedinPACE was associated with poorer neuropsychological test results but provided negligible benefit to clinical predictions.

To our best knowledge, this is the first study to test the combined predictive value of MA, BA and FI in prediction of dementia-related outcomes. Classification performance for normal cognition versus MCI was 0.82, and 0.83 for MCI versus DEM in models including both BA and FI - suggesting good discriminatory performance for these challenging diagnostic tasks. Our best model for distinguishing those with normal cognition from DEM included age, sex and FI, achieving an AUC of 0.95 in the held-out test set, indicating excellent classification. The results confirm the significant relationship between indices of frailty and cognitive impairment previously found in meta-analysis [4]. While both frailty and DEM diagnosis are based on evaluation of function––and thus may overlap–– previous research shows that level of frailty is still predictive for DEM risk independently of global cognition (e.g., CDR-SB) and neuropsychological test performance (e.g., MMSE) [6, 35, 42]. As such, our results strengthen the notion that frailty should be integral in the assessment of older individuals seeking evaluation for cognitive problems [2].

Studies examining both phenotypic and functional-level aging biomarkers for predicting individuals’ level of cognitive impairment are scarce. Our results are in line, however, with the landmark investigation of postmortem neuropathology and frailty by Wallace, et al. [39]. Here, the authors investigated the predictive value of a 41-item FI and neuropathological index (NI) counting the number of diverse pathologies on postmortem brain examination on the prediction of normal cognition, MCI and DEM. Both indices significantly classified CN versus MCI (in-sample AUCs of 0.64 and 0.58 for NI and FI, respectively) and MCI versus DEM (in-sample AUCs of 0.70 and 0.68, respectively). For the same classifications, we obtained similar or superior out-of-sample AUCs using BA and FI, which confirms the predictive value of FI and highlights BA as a putative non-invasive brain marker for clinical diagnosis [29]. The unique contribution of aBA in prediction of 5-year DEM risk reported here, on top of frailty and conventional biomarkers, is novel and strengthens a multidimensional view of DEM [39].

Our findings of sex differences in the prognostic impact of BA and FI on 5-year DEM risk corroborate recent work by Phyo et al. [30]. We found that increased BA had a more adverse impact on 5-year prognosis in females with MCI than males, although this effect was attenuated when adjusting for APOE4 and cognitive tests. Conversely, increased frailty had a more negative effect on prognosis in males, aligning with the well-documented "sex-frailty paradox" where frailty has a greater adverse impact on mortality risk in males [13, 41]. The stronger adverse impact of frailty on DEM risk in males is, however, novel (c.fr. [41]) and merits further study.

Our findings suggest that higher-level biological aging biomarkers, which theoretically represent more advanced stages of aging, are more strongly associated with DEM risk. Specifically, in all comparisons, FI outperformed BA in predictive performance.

Furthermore, when predicting 5-year DEM progression, the coefficients of aFI and aBA were 0.69 and 0.43 respectively, hinting towards the predictive superiority of the former. MA provided negligible predictive value when included alongside BA or FI. While novel in the context of DEM, the results align with one study of mortality risk. Kim et al. compared a 34- item FI with MA in predicting 4.4-year mortality risk using Cox regressions [17]. Individually, MA strongly predicted mortality, but the relationship became non-significant when accounting for baseline frailty. Conversely, Li et al. found GrimAge, PhenoAge, and a mortality risk score DNAm algorithm predicted 17-year mortality risk independently of a 34- item FI [23]. Also, while Cole, et al. [5], found that combining measures of MA and BA improved mortality risk prediction, we failed to find any added predictive power of combining the two in prediction of dementia-related outcomes (Supplemental Fig. 2). The discrepancy in predictive power of the aging biomarker in classifying mortality and dementia-related outcomes needs further investigation.

To our knowledge, no prior studies have evaluated both FI and DunedinPACE for DEM risk. Discrepancies in our findings compared to previous reports might be partially explained by differences in follow-up duration. For example, Thomas et al., reported a 34% increased DEM risk over 14 years for each 1-SD increase in baseline DunedinPACE with Kaplan-Meier curves showing divergence in risk for all DunedinPACE tertiles from 6 to 7 years onwards [37]. These results, along with those of Li et al. [22], suggest the intriguing possibility that different aging biomarkers might play temporally distinct roles in risk prediction across the lifespan, supporting a life course model of DEM risk.

We found an association between DunedinPACE and neuropsychological test results, aligning with Sugden et al., [36]. The results suggest age acceleration (as reflected by the chronological age-adjusted DunedinPACE score) may relate to the biological underpinnings of variation in cognitive function. Unlike Sugden et al., [36], we did not find group-level differences in DunedinPACE between cognitive status levels (CN, MCI, DEM) in our study which uses the same ADNI data. We believe the discrepancy is due to our study’s smaller, split sample and exclusion criteria, resulting in reduced statistical power.

## Limitations

Our study has limitations. Firstly, our sample size was relatively small and the nature of sampling in ADNI may limit generalizability to the broader population. Replication in larger, population-level samples such as UK Biobank would be valuable. Furthermore, the sample size was particularly small with respect to participants with methylation data, which could explain the lack of predictability deduced from the MA measures. Secondly and relatedly, we examined molecular aging using a limited set of published MA algorithms. As the field progresses, incorporating novel MA measures may offer better predictive value than what we report here. Thirdly, we focused on dementia-related *functional* outcome measures (i.e., clinical diagnosis), not disease-pathology measures such as amyloid or tau. Future studies may evaluate the interplay between aging biomarkers, disease-specific markers and cognition for a more complete understanding of DEM development and risk (see, e.g., [25]). Finally, we used single-timepoint measurement of the aging biomarkers, which limits interpretation of temporal dynamics of aging reflected by the markers. For instance, intriguing research by Vidal-Pineiro, et al., has shown that BA variation may reflect static, early life factors above current rates of brain aging [38]. While this caveat pertaining to all cross-sectional data is central to the interpretation of the biological underpinnings of the biomarkers, it does not halt their practical use in predictive medicine.

## Conclusion

Our results underscore the potential predictive value of combining higher-level biological aging biomarkers in the context of dementia-related outcomes. Future research could explore the biomarkers longitudinally and across longer time frames, tracking changes within individuals to pinpoint inflection points in the aging process and uncover stages of the life span where they provide most value in risk assessment. The results suggest caution in using or marketing MA algorithms such as DunedinPACE as individual risk markers for shorter-term prognosis or prediction of current cognitive impairment. Additionally, studies integrating disease-specific markers (amyloid, tau) with the present aging-related biomarkers could further our understanding of the interplay between aging and disease.

## Statements and Declarations

## Data Availability

Data are available at ADNI website. Code will be shared upon request.

## Acknowledgements

Data collection and sharing for ADNI is funded by the National Institute on Aging (National Institutes of Health Grant U19 AG024904). The grantee organization is the Northern California Institute for Research and Education. In the past, ADNI has also received funding from the National Institute of Biomedical Imaging and Bioengineering, the Canadian Institutes of Health Research, and private sector contributions through the Foundation for the National Institutes of Health (FNIH) including generous contributions from the following: AbbVie, Alzheimer’s Association; Alzheimer’s Drug Discovery Foundation; Araclon Biotech; BioClinica, Inc.; Biogen; Bristol-Myers Squibb Company; CereSpir, Inc.; Cogstate; Eisai Inc.; Elan Pharmaceuticals, Inc.; Eli Lilly and Company; EuroImmun; F. Hoffmann-La Roche Ltd and its affiliated company Genentech, Inc.; Fujirebio; GE Healthcare; IXICO Ltd.; Janssen Alzheimer Immunotherapy Research & Development, LLC.; Johnson & Johnson Pharmaceutical Research &Development LLC.; Lumosity; Lundbeck; Merck & Co., Inc.; Meso Scale Diagnostics, LLC.; NeuroRx Research; Neurotrack Technologies; Novartis Pharmaceuticals Corporation; Pfizer Inc.; Piramal Imaging; Servier; Takeda Pharmaceutical Company; and Transition Therapeutics. We acknowledge Luigi A. Maglanoc, Ph.D., for his instrumental role in constructing the FI used in the analyses.

