## Supplementary Information for "Complementary value of molecular, phenotypic and functional aging biomarkers in dementia prediction"

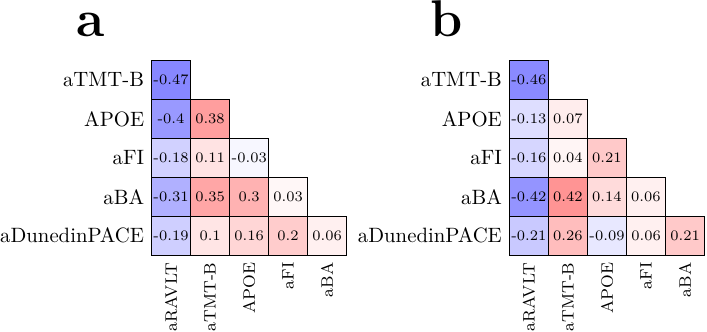

Supplementary Fig. 1 The figure shows Pearson correlations among the putative aging and clinical biomarkers for a) females, and b) males. Abbreviations: prefix "a" means that the standardized age- and sex-adjusted residual of the biomarker is used as input in the correlation analysis. APOE = Number of apolipoprotein E epsilon 4 alleles (0, 1 or 2), BA = Brain age, FI = Frailty index, RAVLT = Rey Auditory Verbal Learning Test, immediate recall, TMT-B = Trail Making Test, part B

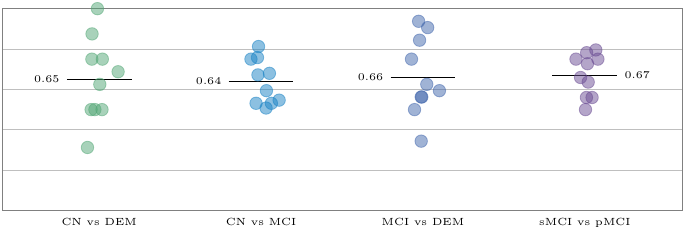

Supplementary Fig. 2 Predictive performance––reported as Area under the Receiver Operating Characteristic Curve (AUC)–– for classification models including age- and sex-adjusted brain age (aBA) and aDunedinPACE as predictors. Three models classified baseline cognitive status: CN = cognitively normal, DEM = dementia, MCI = mild cognitive impairment. One model predicted 5-year MCI conversion to DEM, comparing stable (sMCI) vs. progression (pMCI). Age and sex were used as covariates in all models. Individual points represent performance (AUC) in independent folds in cross-validation, whereas the black line denotes their mean
